# Clinical, molecular, and immune correlates of *Immunotherapy Response Score* in advanced urothelial carcinoma patients under atezolizumab monotherapy: Analysis of the phase 2 IMvigor210 trial

**DOI:** 10.1101/2023.04.11.23288423

**Authors:** Miriam Ferreiro-Pantín, Urbano Anido-Herranz, Yoel Z. Betancor, Víctor Cebey-López, Luis León-Mateos, Jorge García-González, Silvia Margarita García-Acuña, Natalia Fernández-Díaz, Jose M. C. Tubio, Rafael López-López, Juan Ruiz-Bañobre

## Abstract

**Background:** In the advanced urothelial carcinoma (aUC) scenario there are no consistent biomarkers to predict the benefit patients derived from immune checkpoint blockade. Recently a novel pan-tumor molecular tissue-based biomarker, the Immunotherapy Response Score (IRS), has been proposed. Herein we conducted a retrospective study to validate the prognostic and predictive utility of the IRS in aUC patients under atezolizumab monotherapy and to characterize its underline molecular and immune features in the context of the IMvigor210 phase 2 clinical trial.

**Methods:** This is a retrospective study of 261 patients with available clinical, molecular, and immune tumor data treated with atezolizumab monotherapy in the IMvigor210 phase 2 clinical trial. Efficacy endpoints were overall survival (OS), disease control rate (DCR), and overall response rate (ORR). Survival estimates were calculated by the Kaplan Meier method, and groups were compared with the log-rank test. The Cox proportional hazards regression model was used to evaluate factors independently associated with OS. Factors associated with disease control (DC) and response were tested with logistic regression in univariable and multivariable analyses. Comparisons between patient and disease characteristics were carried out using Chi-squared or Fisher exact tests. All p values were 2-sided, and those less than 0.05 were considered statistically significant.

**Results:** High IRS was significantly associated with a better OS in univariable [hazard ratio (HR)=0.49, 95% CI 0.33–0.74, p<0.001] and multivariable (HR=0.57, 95% CI 0.37–0.86, p=0.007) analyses. DCR and ORR were significantly higher among high IRS patients (DCR for high IRS vs low IRS patients: 57% vs 32%, p<0.001; ORR for high IRS vs low IRS patients: 42% vs 10%, p<0.001). High IRS patients presented a higher probability of DC and response in univariable [DC: odds ratio (OR)=2.72, 95% CI 1.54–4.81, p<0.001; Response: OR=3.92, 95% CI 2.11–7.31; p<0.001] and multivariable (DC: OR=2.38, 95% CI 1.28–4.44, p=0.006; Response: OR=3.36, 95% CI 1.68–6.69, p<0.001) analyses.

**Conclusions:** This study validates IRS as a strong independent prognostic and predictive biomarker for OS and DC/response in aUC patients treated with atezolizumab monotherapy in the IMvigor210 phase 2 clinical trial.

**Clinical Trial Registration:** NCT02108652, NCT02951767.

## Introduction

Bladder cancer is the tenth most commonly diagnosed cancer worldwide, with approximately 573,278 new cases and 212,536 estimated cancer deaths in 2020^1^. Though immunotherapy, particularly programmed cell death-1 (PD-1) and programmed death-ligand 1 (PD-L1) blockers, has revolutionized cancer management in recent years, the combination platinum-based chemotherapy is still the standard of care for first-line treatment of advanced urothelial carcinoma (aUC)^2–4^. Today, the use of avelumab, an anti-PD-L1 antibody, is indicated as first-line maintenance therapy if the disease has not progressed on platinum-based chemotherapy. Among those patients whose disease has progressed on a previous platinum-based strategy, pembrolizumab (an anti-PD-1 antibody) or atezolizumab (an anti-PD-L1 antibody) are the recommended treatment options^3,4^. Upfront therapy with single-agent immune checkpoint blockade (ICB), either pembrolizumab or atezolizumab, is indicated only for those patients who are ineligible for platinum-based treatment^3,4^.

Over the last decade, a plethora of studies have evaluated the role of different prognostic and/or predictive biomarkers for ICB in aUC. Numerous translational research initiatives have explored the role of different molecular markers such as PD-L1^5^, tumor mutational burden (TMB)^6,7^, copy-number and single-nucleotide variant counts^8^, alterations in DNA damage response and repair genes^9^, gene expression signatures^10–13^, peripheral blood T-cell receptor clonality^14^, and clinical variables^15^. Despite these huge efforts, to date, there is not any consistent biomarker translated to the clinic. In this regard, Tomlins et al^16^ have recently developed and validated a novel pan-tumor tissue-based biomarker, the Immunotherapy Response Score (IRS), which integrating TMB and the expression of certain genes such as *PD-1, PD-L1, TOP2A*, and *ADAM12* in a Cox model, identifies those patients who derived a higher benefit in terms of time to next therapy [which the authors defined as real world progression-free survival (rwPFS)] and overall survival (OS) when treated with single-agent anti-PD-1 or anti-PD-L1 immunotherapy. However, the correlation of IRS with other important clinical outcomes such as disease control or response was not evaluated. Taking this into consideration, herein we conducted a retrospective study in order to validate the prognostic and predictive role of IRS in patients diagnosed with aUC treated with atezolizumab in the context of the IMvigor210 phase 2 clinical trial^5,17^. Additionally, we explored the correlation of IRS with different molecular and immune tumor characteristics.

## Patients and Methods

### Study design and patient population

The design and primary outcomes of the single-arm phase 2 study of atezolizumab in aUC (IMvigor210) were described in previous reports^5,17^. This is a retrospective study of 261 patients with available clinical, molecular, and immune tumor data from the IMvigor210^10^. For the purpose of our analyses, our efficacy endpoints were overall survival (OS), disease control rate (DCR), and overall response rate (ORR). Tumor responses were assessed according to Response Evaluation Criteria in Solid Tumors guidelines version 1.1^5^.

Individual patient IRS were derived from the Cox model as previously described (IRS = 0.273758 * TMB + 0.112641 * *PD-1* + 0.061904 * *PD-L1* - 0.077011 * *TOP2A* - 0.057991 * *ADAM12*) and considered as a binary predictor based on the previously defined cut-off threshold (high ≥0.873569 vs low <0.873569)^16^. Tumor mutational burden was calculated as mutations per megabase (Mb) of genomic target territory of the FoundationOne panel^10^. TMB-high patients were defined as those with a TMB ≥10 mutations/Mb. Whole transcriptome profiles were generated using TruSeq RNA Access technology (Illumina)^10^. Raw count data for the genes of interest were transformed to log_2_ normalized reads per million (RPM), and values for each gene were median centered across a representative reference clinical population, The Cancer Genome Atlas Urothelial Bladder Carcinoma cohort. Nine samples with normalized RPM equal to 0 for any of the 4 genes were removed. Differential gene expression analysis was performed with the R package *DeSeq2* version 1.36.0. Gene set enrichment analysis (GSEA) was performed with the R package *clusterProfiler* version 4.6.2.

Statements confirming compliance with ethical regulations, the committees that approved the IMvigor210 study protocol, and confirmation of informed consent from all study participants are included in the previous publications describing the IMvigor210 trial (NCT02108652)^5,10,17^.

### Statistical analysis

Survival estimates were calculated by the Kaplan Meier method, and groups were compared with the log-rank test. The Cox proportional hazards regression model was used to evaluate factors independently associated with overall survival (OS). Baseline variables included in the multivariable analysis were selected according to statistical significance in univariable analysis (cutoff, p value <0.05). The proportional hazard assumption was verified with the Schoenfeld residual method. Factors associated with disease control (DC) and response were tested with logistic regression in univariable analyses. Variables included in the final multivariable model were selected according to their statistical significance in univariable analysis (cutoff, p value <0.05). Comparisons between patient and disease characteristics were carried out using Chi-squared or Fisher exact tests. All p values were 2-sided, and those less than 0.05 were considered statistically significant. The Benjamini–Hochberg procedure was used to control the false discovery rate in case of multiple comparisons. All statistical analyses were performed using R version 4.2.2 (Vienna, Austria).

## Results

### Patient population

From 348 patients enrolled in the IMvigor210 trial and treated with atezolizumab, 261 had all the clinical, molecular, and immune tumor data to be included in this retrospective study (**Supplementary Figure 1**). Baseline patient and disease characteristics are summarized in **Table 1**. Twenty six percent (n=67) of patients had a high IRS, while 74% (n=194) had low IRS. The distribution of different patient and disease characteristics according to IRS is shown in **Table 1**. To note, in the IRS high group there was a higher proportion of patients with genomically unstable Lund taxonomy subtype (p=0.002), PD-L1 expression on tumor-infiltrating immune cells (IC) ≥5% (IC2/3) (p<0.001), PD-L1 expression on tumor cells (TC) ≥5% (TC2/3) (p=0.040), and immune inflamed phenotype (p<0.001). Importantly, there were no statistically significant differences in TMB (high vs low) distribution among IRS high and IRS low cases.

**Table 1.** Distribution of IRS according to patient and disease characteristics.

| Characteristics | Total (n = 261) | IRS high (n = 67, 26%) | IRS low (n = 194, 74%) | p value |
| --- | --- | --- | --- | --- |
| **Sex - no. (%)** |  |  |  | 0.072 |
| Female | 53 (20) | 8 (12) | 45 (23) |  |
| Male | 208 (80) | 59 (88) | 149 (77) |  |
| **Previous intravesical BCG - no. (%)** |  |  |  | 0.766 |
| No | 204 (78) | 51 (76) | 153 (79) |  |
| Yes | 57 (22) | 16 (24) | 41 (21) |  |
| **Smoking status - no. (%)** |  |  |  | 0.093 |
| Never | 86 (33) | 16 (24) | 70 (36) |  |
| Ever | 175 (67) | 51 (76) | 124 (64) |  |
| **Previous platinum-based therapy - no. (%)** |  |  |  | 0.865 |
| No | 70 (27) | 19 (28) | 51 (26) |  |
| Yes | 191 (73) | 48 (78) | 143 (74) |  |
| **AJCC stage at diagnosis - no. (%)** |  |  |  | 0.838 |
| I - II | 155 (59) | 41 (61) | 114 (59) |  |
| III - IV | 106 (41) | 26 (39) | 80 (41) |  |
| **IC PD-L1 level - no. (%)** |  |  |  | **<0.001** |
| IC0/1 | 159 (61) | 27 (40) | 132 (68) |  |
| IC2/3 | 102 (39) | 40 (60) | 62 (32) |  |
| **Immune phenotype - no. (%)** |  |  |  | **<0.001** |
| Inflamed | 170 (75) | 55 (95) | 115 (68) |  |
| Non-inflamed | 57 (25) | 3 (5) | 54 (32) |  |
| **TC PD-L1 level - no. (%)** |  |  |  | **0.040** |
| TC0/1 | 221 (85) | 51 (76) | 170 (88) |  |
| TC2/3 | 40 (15) | 16 (24) | 24 (12) |  |
| **ECOG-PS - no. (%)** |  |  |  | 0.285 |
| 0 | 102 (39) | 22 (33) | 80 (41) |  |
| 1 | 159 (61) | 45 (67) | 114 (59) |  |
| **TMB - no. (%)** |  |  |  | 0.069 |
| High | 10 (4) | 67 (100) | 184 (95) |  |
| Low | 251 (96) | 0 (0) | 10 (5) |  |
| **Lund taxonomy - no. (%)** |  |  |  | **0.002** |
| Genomically unstable | 59 (23) | 25 (37) | 34 (18) |  |
| Non-genomically unstable | 202 (77) | 42 (63) | 160 (82) |  |

Bold numbers indicate statistically significant values.
Abbreviations: AJCC, American Joint Committee on Cancer; BCG, Bacillus Calmette-Guérin; CI, confidence interval; ECOG-PS, Eastern Cooperative Oncology Group performance status; HR, hazard ratio; IC, tumor-infiltrating immune cells; IRS, Immunotherapy Response Score; PD-L1, programmed death-ligand 1; TC, tumor cells; TMB, tumor mutational burden.

### Clinical significance of Immunotherapy Response Score

#### Overall Survival

Among 261 cases included in this retrospective study, median OS was 8.90 months (95% CI 7.06–10.91) (**Supplementary Table 1**). Median OS for high and low IRS patients was 16.46 months (95% CI 10.58–17.28) and 7.43 months (95% CI 5.85–9.56) (p<0.001), respectively (**Figure 1**). High IRS was significantly associated with a better OS in univariable [hazard ratio (HR)=0.49, 95% CI 0.33–0.74, p<0.001] and multivariable (HR=0.57, 95% CI 0.37– 0.86, p=0.007) analyses (**Table 2**). Other baseline variables independently associated with a better OS in multivariable analysis were ECOG-PS 0 (HR=0.40, 95% CI 0.28–0.56, p<0.001), genomically unstable Lund taxonomy subtype (HR=0.46, 95% CI 0.30–0.71, p<0.001), and PD-L1 expression on tumor-infiltrating immune cells (IC) ≥5% (IC2/3) (HR=0.67, 95% CI 0.48–0.94, p=0.020) (**Table 2**). High TMB was not associated with an improved OS (**Table 2**).

**Table 2.** Univariable and multivariable Cox regression analyses for overall survival.

| Characteristics | Univariable analysis |  | Multivariable analysis |  |
| --- | --- | --- | --- | --- |
|  | HR (95% CI) | p value | HR (95% CI) | p value |
| Sex (male vs female) | 0.95 (0.66 - 1.38) | 0.927 |  |  |
| Previous intravesical BCG (yes vs no) | 1.03 (0.72 – 1.48) | 0.927 |  |  |
| Smoking status (ever vs never) | 0.91 (0.66 – 1.25) | 0.894 |  |  |
| Previous platinum-based therapy (yes vs no) | 1.21 (0.84 – 1.73) | 0.559 |  |  |
| AJCC stage at diagnosis (III – IV vs I – II) | 1.21 (0.89 – 1.64) | 0.498 |  |  |
| IC PD-L1 level (IC2/3 vs IC0/1) | 0.59 (0.42 – 0.81) | **0.005** | 0.67 (0.48 – 0.94) | **0.020** |
| Immune phenotype (inflamed vs non-inflamed) | 0.71 (0.49 – 1.01) | 0.148 |  |  |
| TC PD-L1 level (TC2/3 vs TC0/1) | 0.98 (0.65 – 1.49) | 0.927 |  |  |
| ECOG-PS (0 vs 1) | 0.46 (0.33 – 0.64) | **<0.001** | 0.40 (0.28 – 0.56) | **<0.001** |
| TMB (high vs low) | 0.94 (0.44 – 2.00) | 0.927 |  |  |
| Lund taxonomy (genomically unstable vs non-genomically unstable) | 0.47 (0.31 – 0.72) | **0.002** | 0.46 (0.30 – 0.71) | **<0.001** |
| IRS (high vs low) | 0.49 (0.33 – 0.74) | **<0.001** | 0.57 (0.37 – 0.86) | **0.007** |

Bold numbers indicate statistically significant values.
Abbreviations: AJCC, American Joint Committee on Cancer; BCG, Bacillus Calmette-Guérin; CI, confidence interval; ECOG-PS, Eastern Cooperative Oncology Group performance status; HR, hazard ratio; IC, tumor-infiltrating immune cells; IRS, Immunotherapy Response Score; PD-L1, programmed death-ligand 1; TC, tumor cells; TMB, tumor mutational burden.

**Figure 1.**
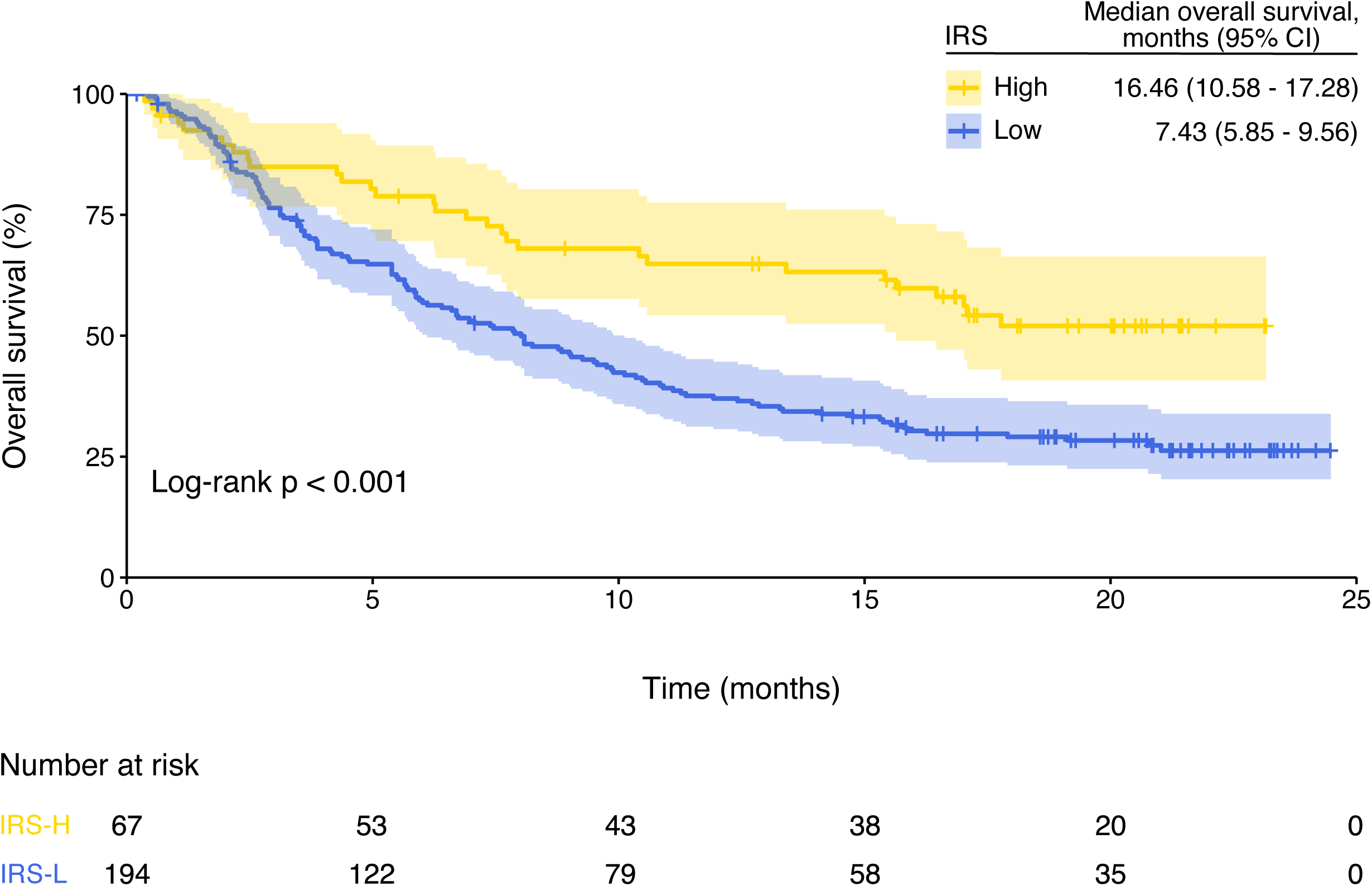
Kaplan–Meier overall survival estimates according to Immunotherapy Response Score (IRS). Abbreviations: IRS high, IRS-H; IRS low, IRS-L.

#### Disease control and response

Among 261 cases included in this retrospective study, disease control rate (DCR) and overall response rate (ORR) were 38.70% (95% CI 32.75–44.90%) and 22.22% (95% CI 17.33–27.56%) respectively, including 21 (8.05%) complete responses (**Supplementary Table 1**). DCR and ORR were significantly higher among high IRS patients (DCR for high IRS vs low IRS patients: 57% vs 32%, p<0.001; ORR for high IRS vs low IRS patients: 42% vs 10%, p<0.001) (**Figure 2**). High IRS patients presented a higher probability of DC and response in univariable analysis [DC: odds ratio (OR)=2.72, 95% CI 1.54–4.81, p<0.001; Response: OR = 3.92, 95% CI 2.11–7.31; p<0.001]. Other variables associated with a higher probability of disease control and response in univariable analysis were ECOG-PS 0 (DC: OR=2.04, 95% CI 1.22–3.40, p=0.006; Response: OR=1.95, 95% CI 1.08–3.52; p=0.027), PD-L1 expression on tumor-infiltrating IC2/3 (DC: OR=2.04, 95% CI 1.22–3.40, p=0.006; Response: OR=2.13, 95% CI 1.18– 3.86; p=0.012), and genomically unstable Lund taxonomy subtype (DC: OR=2.50, 95% CI 1.39– 4.52, p=0.002; Response: OR=3.39, 95% CI 1.79–6.41; p<0.001) (**Table 3** and **Table 4**). When these variables were evaluated in multivariable analysis, IRS, ECOG-PS, and Lund taxonomy were independently associated with a higher probability of DC (high IRS: OR=2.38, 95% CI 1.28–4.44, p=0.006; ECOG-PS 0: OR=2.20, 95% CI 1.27–3.79, p=0.005; genomically unstable Lund taxonomy subtype: OR=2.42, 95% CI 1.28–4.56, p=0.006) and response (high IRS: OR=3.36, 95% CI 1.68– 6.69, p<0.001; ECOG-PS 0: OR=2.28, 95% CI 1.18–4.37, p=0.014; genomically unstable Lund taxonomy subtype: OR=3.11, 95% CI 1.55–6.23, p=0.001) (**Table 3** and **Table 4**).

**Figure 2.**
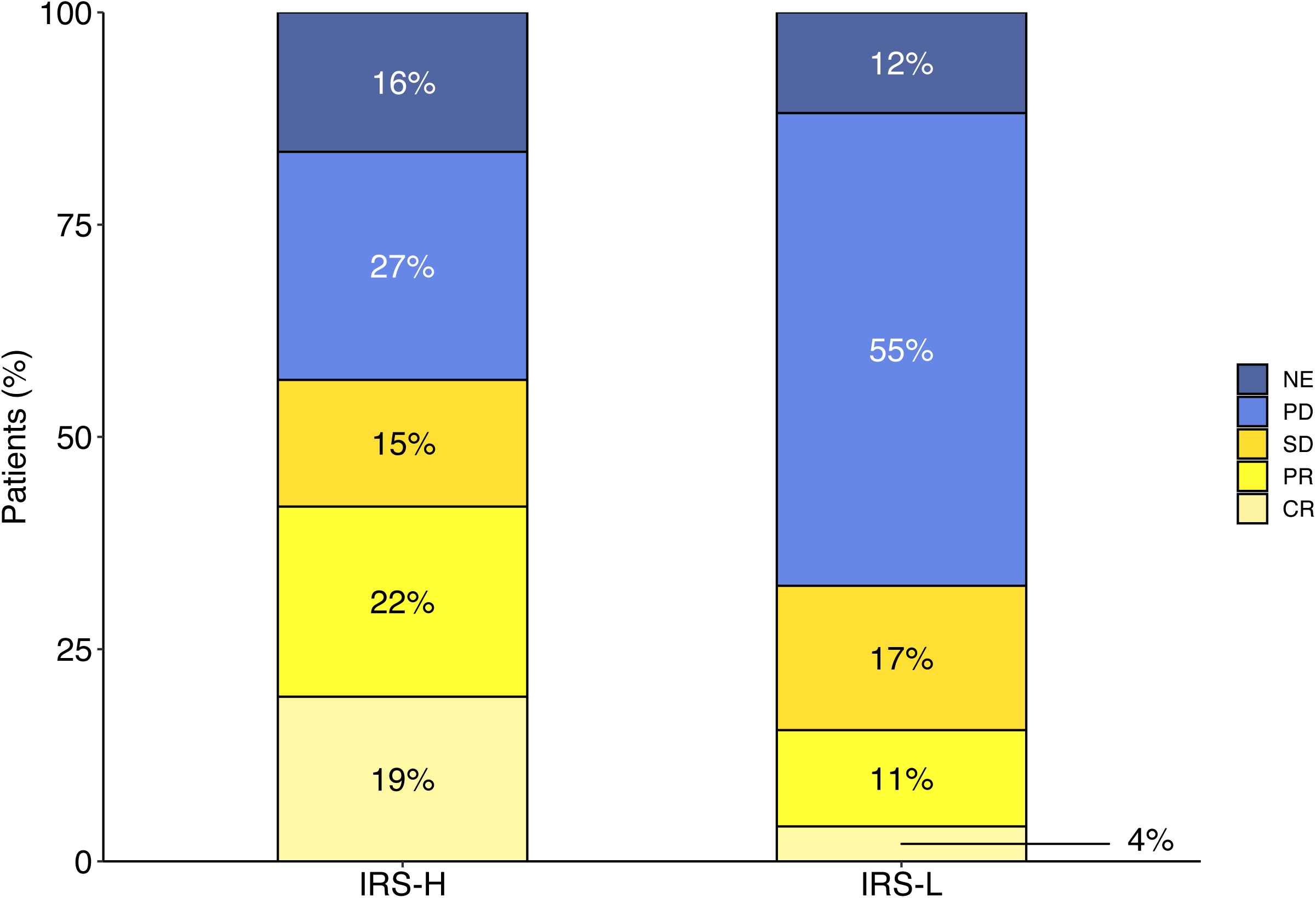
Atezolizumab response distribution by Immunotherapy Response Score (IRS). NE, not evaluable; PD, progressive disease; SD, stable disease; PR, partial response; CR, complete response.

**Table 3.** Univariable and multivariable logistic regression analyses for disease control.

| Characteristics | Univariable analysis |  | Multivariable analysis |  |
| --- | --- | --- | --- | --- |
|  | OR (95% CI) | p value | OR (95% CI) | p value |
| Sex (male vs female) | 0.86 (0.47 – 1.59) | 0.638 |  |  |
| Previous intravesical BCG (yes vs no) | 1.00 (0.54 – 1.82) | 0.986 |  |  |
| Smoking status (ever vs never) | 1.48 (0.86 – 2.55) | 0.154 |  |  |
| Previous platinum-based therapy (yes vs no) | 0.73 (0.42 – 1.27) | 0.263 |  |  |
| AJCC stage at diagnosis (III – IV vs I – II) | 0.87 (0.52 – 1.45) | 0.601 |  |  |
| IC PD-L1 level (IC2/3 vs IC0/1) | 2.04 (1.22 – 3.40) | **0.006** | 1.66 (0.95 – 2.89) | 0.074 |
| Immune phenotype (inflamed vs non-inflamed) | 1.63 (0.86 – 3.08) | 0.132 |  |  |
| TC PD-L1 level (TC2/3 vs TC0/1) | 1.36 (0.69 – 2.68) | 0.375 |  |  |
| ECOG-PS (0 vs 1) | 2.04 (1.22 – 3.40) | **0.006** | 2.32 (1.34 – 4.03) | **0.003** |
| TMB (high vs low) | 0.94 (0.26 – 3.43) | 0.931 |  |  |
| Lund taxonomy (genomically unstable vs non-genomically unstable) | 2.50 (1.39 – 4.52) | **0.002** | 2.42 (1.28 – 4.56) | **0.006** |
| IRS (high vs low) | 2.72 (1.54 – 4.81) | **<0.001** | 2.33 (1.25 – 4.35) | **0.008** |

Bold numbers indicate statistically significant values.
Abbreviations: AJCC, American Joint Committee on Cancer; BCG, Bacillus Calmette-Guérin; CI, confidence interval; ECOG-PS, Eastern Cooperative Oncology Group performance status; HR, hazard ratio; IC, tumor-infiltrating immune cells; IRS, Immunotherapy Response Score; PD-L1, programmed death-ligand 1; TC, tumor cells; TMB, tumor mutational burden.

**Table 4.** Univariable and multivariable logistic regression analyses for response.

| Characteristics | Univariable analysis |  | Multivariable analysis |  |
| --- | --- | --- | --- | --- |
|  | OR (95% CI) | p value | OR (95% CI) | p value |
| Sex (male vs female) | 1.11 (0.53 – 2.33) | 0.774 |  |  |
| Previous intravesical BCG (yes vs no) | 1.18 (0.59 – 2.36) | 0.631 |  |  |
| Smoking status (ever vs never) | 1.24 (0.66 – 2.34) | 0.504 |  |  |
| Previous platinum-based therapy (yes vs no) | 0.77 (0.40 – 1.45) | 0.412 |  |  |
| AJCC stage at diagnosis (III – IV vs I – II) | 0.72 (0.39 – 1.32) | 0.282 |  |  |
| IC PD-L1 level (IC2/3 vs IC0/1) | 2.13 (1.18 – 3.86) | **0.012** | 1.58 (0.82 – 3.06) | 0.176 |
| Immune phenotype (inflamed vs non-inflamed) | 1.54 (0.72 – 3.32) | 0.268 |  |  |
| TC PD-L1 level (TC2/3 vs TC0/1) | 0.86 (0.37 – 1.97) | 0.714 |  |  |
| ECOG-PS (0 vs 1) | 1.95 (1.08 – 3.52) | **0.027** | 2.38 (1.23 – 4.60) | **0.010** |
| TMB (high vs low) | 2.64 (0.33 – 21.31) | 0.361 |  |  |
| Lund taxonomy (genomically unstable vs non-genomically unstable) | 3.39 (1.79 – 6.41) | **<0.001** | 3.11 (1.55 – 6.23) | **0.001** |
| IRS (high vs low) | 3.92 (2.11 – 7.31) | **<0.001** | 3.28 (1.64 – 6.56) | **<0.001** |

Bold numbers indicate statistically significant values.
Abbreviations: AJCC, American Joint Committee on Cancer; BCG, Bacillus Calmette-Guérin; CI, confidence interval; ECOG-PS, Eastern Cooperative Oncology Group performance status; HR, hazard ratio; IC, tumor-infiltrating immune cells; IRS, Immunotherapy Response Score; PD-L1, programmed death-ligand 1; TC, tumor cells; TMB, tumor mutational burden.

### Biological significance of the Immunotherapy Response Score

To fully characterize IRS from a biological viewpoint, we carried out a differential gene expression analysis (**Figure 3A**) followed by a GSEA (**Figure 3B, Supplementary Figure 2A,B**, and **Supplementary Table 2A-C**). As expected, this analysis revealed an enrichment of important biological processes associated with immune system activation such as natural killer cell mediated immunity (adjusted p value<0.001), lymphocyte mediated immunity (adjusted p value<0.001), lymphocyte migration (adjusted p value<0.001), and response to interferon-gamma (adjusted p value<0.001) in IRS high cases. On the contrary, processes associated with stroma such as extracellular matrix organization (adjusted p value<0.001) and extracellular structure organization (adjusted p value<0.001) were up regulated in IRS low cases (**Figure 3B**). Other statistically significantly enriched Gene Ontology components (Molecular Function and Cellular Component) are described in **Supplementary Figure 2A,B** and **Supplementary Table 2A-C**.

**Figure 3.**
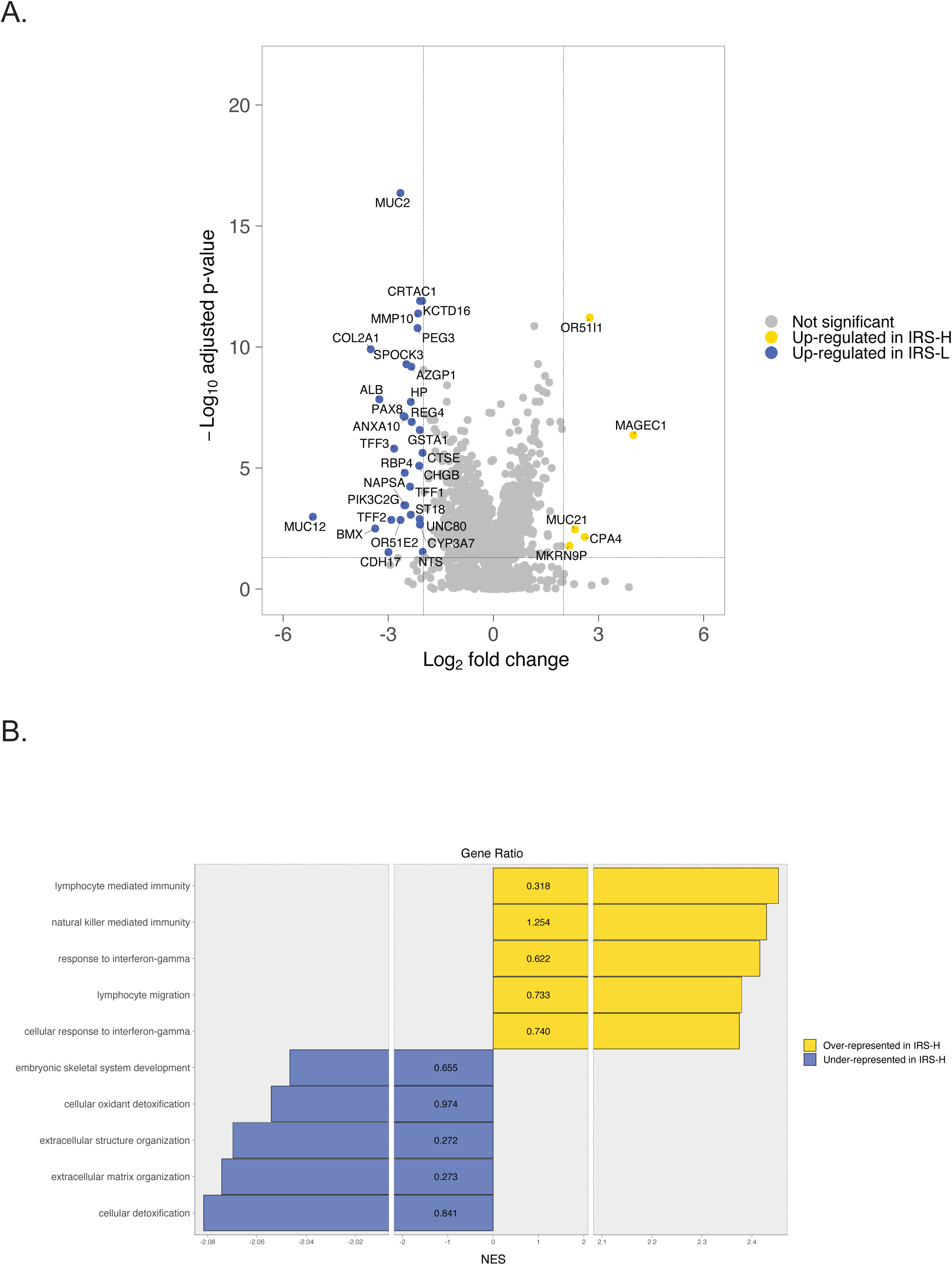
**(A)** Volcano plot representing gene expression differences between Immunotherapy Response Score (IRS) high (IRS-H) and IRS low (IRS-L) cases. **(B)** Gene set enrichment analysis showing statistically significantly over- or under-represented Gene Ontology Biological Processes in IRS-H cases compared with IRS-low cases. For simplicity, biological processes *natural killer cell mediated cytotoxicity, natural killer cell mediated immunity, natural killer cell activation*, and *regulation of natural killer cell mediated cytotoxicity* were represented together under the term *natural killer cell mediated immunity*. In this particular case, represented NES and Gene Ratio are the median of these 4 biological processes.

## Discussion

Multiple studies have been conducted to discover predictive biomarkers for cancer immunotherapy, but to date, only microsatellite instability has been adopted in the clinic as the first tissue/site-agnostic predictive biomarker for the anti-PD-1 antibody pembrolizumab. Though TMB has been also FDA-approved as a predictive biomarker for the same drug in a tissue/site-agnostic cancer indication, its utility in the daily clinical practice is still debatable. Taking this into consideration, Tomlins et al^16^ have developed a new pan-solid tumor prognostic/predictive biomarker, the IRS, which integrating TMB and the normalized expression of *PD-1, PD-L1, TOP2A*, and *ADAM12* genes in a Cox model, identifies those patients who derived a higher benefit in terms of rwPFS and OS when treated with single-agent anti-PD-1 or anti-PD-L1 immunotherapy. Considering the importance of validating biomarkers in prospective cohorts, herein we conducted a retrospective study in order to validate the prognostic and predictive role of IRS in patients diagnosed with aUC treated with atezolizumab in the IMvigor210 phase 2 clinical trial^5^.

First, according to the clinicopathological and tumor molecular features, we found an enrichment of different characteristics classically correlated with more immunogenic tumors in IRS high cases such as the genomically unstable Lund taxonomy subtype, the expression of PD-L1 on tumor-infiltrating IC2/3, the expression of PD-L1 on TC2/3, and the immune inflamed phenotype^18,19^. Interestingly and in line with the work of Tomlins et al^16^, we did not find any statistically significant difference in the TMB status distribution among IRS groups.

Second, we evaluate the correlation of IRS with OS of aUC patients treated with atezolizumab. As expected, IRS demonstrated a strong independent prognostic significance, with IRS high cases presenting a 51% reduction in the risk of death compared with IRS low cases. These results are in line with those reported by Tomlins et al^16^, who found a risk of death reduction of 48% and 51% in the discovery and validation pan-tumor cohorts of their study, respectively. It is important to highlight that our study validates for the first time the prognostic utility of IRS in a prospective cohort of 261 aUC patients treated with atezolizumab. This represents an important step in the aUC clinical scenario, taking into consideration that the original study^16^ was not specifically designed to address this question in this specific tumor type, and only included 62 bladder cancer cases treated with different immune checkpoint inhibitors, either pembrolizumab monotherapy (45 patients) in the discovery cohort, or an alternative anti-PD-1/PD-L1 monotherapy (17 patients: 12 treated with atezolizumab, 3 with nivolumab, and 2 with avelumab) in the validation cohort.

Third, while Tomlins et al^16^ validated the predictive nature of the IRS by using various indirect approaches involving rwPFS, herein we directly demonstrated its ability to predict DC and response. In our study, IRS high cases had not only a higher DCR and ORR, but also an increased probability of DC and response compared to those IRS low cases.

Finally, in an attempt to fully characterize IRS from a biological viewpoint, we carried out a GSEA, which as expected, revealed an enrichment of important biological processes associated with immune system activation in IRS high cases. On the contrary, IRS low cases were enriched in biological processes associated with stroma, which agree with previous findings associating a lack of response to atezolizumab in those aUC patients with an immune-excluded tumor microenvironment with a high pan-fibroblast TGF-β response signature^10^.

Our study has two main limitations. The first one is the use of a prospective cohort from a single-arm phase 2 clinical trial. Although we could evaluate the correlation of IRS with DC and response to atezolizumab, due to the lack of a comparator arm of patients treated with an alternative drug, we were limited to carry out a test of interaction to definitively demonstrate the predictive value of IRS in this particular clinical scenario. This point could be clarified validating our results in a prospective cohort from a randomized phase 3 clinical trial. The second limitation is the use of different molecular platforms to estimate the TMB and the expression of genes comprising the IRS. Though originally the IRS includes TMB from a StrataNGS comprehensive genomic profiling test, and expression of *PD-1, PD-L1, TOP2A*, and *ADAM12* genes from a multiplex PCR-based quantitative transcriptional profiling test, in our study TMB and gene expression were evaluated by using a FoundationOne panel and whole transcriptome sequencing, respectively. However, from a pragmatic viewpoint and considering the high concordance demonstrated in a previous validation study between TMB estimated with either the StrataNGS comprehensive genomic profiling assay or the FoundationOne panel^20^, we feel confident about the robustness and interchangeability of our results.

Today, either in daily clinical practice or in a clinical trial scenario, there are available different treatment options for the management of patients with aUC. In this context, the development of tools to help in the decision-making process is mandatory. In this study, in addition to demonstrating the prognostic and predictive utility of the IRS in aUC patients under atezolizumab monotherapy, we characterized its underline molecular and immune features. If the results of this study are definitively validated, the IRS will represent a valuable tool for therapy selection in this setting: immunotherapy yes or not, alone or in combination.

## Data Availability

All data used for the analyses presented here have been previously deposited to the European Genome-Phenome Archive under accession number EGAS00001002556 and is available in IMvigor210CoreBiologies, a fully documented software and data package for the R statistical computing environment. This package is freely available under the Creative Commons 3.0 license and can be downloaded from http://research-pub.gene.com/IMvigor210CoreBiologies/packageVersions/IMvigor210CoreBiologies_1.0.0.tar.gz.

## Acknowledgements

MF-P is supported by a Santander Investigación predoctoral research contract from the University of Santiago de Compostela.

YZB was supported by a Programa Investigo 2022 research contract from the Consellería de Emprego e Igualdade and is supported by a predoctoral fellowship from Xunta de Galicia (ED481A 2022/491).

JR-B is supported by a Juan Rodés contract (JR21/00019) from the Institute of Health Carlos III. The results shown here are in part based upon data generated by the TCGA Research Network: https://www.cancer.gov/tcga.

We thank the Supercomputing Centre of Galicia (CESGA) for providing complementary computational resources.

## Author Contributions

Conceptualization: MF-P and JR-B; software, MF-P and JR-B; validation, MF-P and JR-B; formal analysis, MF-P and JR-B; investigation: all authors; resources: all authors; data curation, all authors; writing – original draft, MF-P and JR-B; writing – review & editing, all authors; visualization, MF-P, YZB, and JR-B; supervision, JR-B; project administration, J.R.-B; funding acquisition, JR-B.

## Figure legends

**Supplementary Figure 1.**
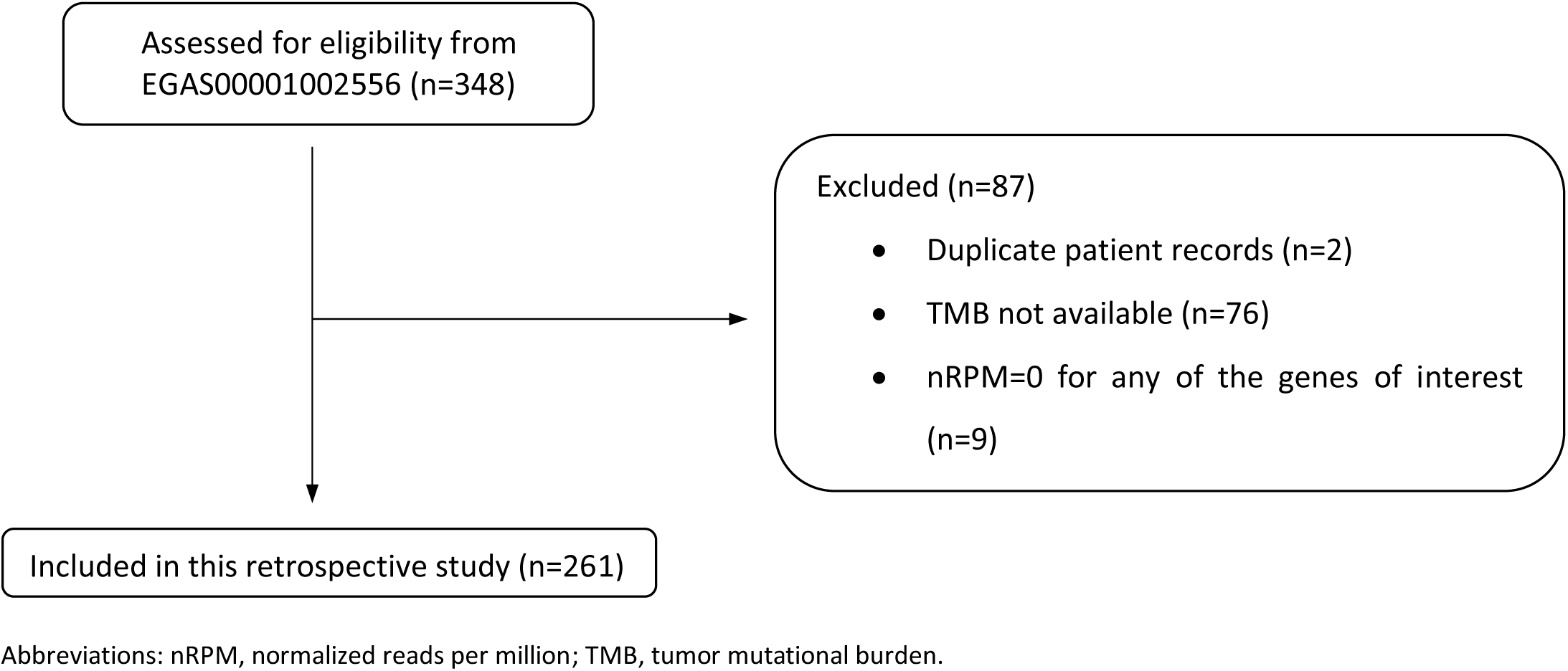
Study flow diagram. Abbreviations: nRPM, normalized reads per million; TMB, tumor mutational burden.

**Supplementary Figure 2.**
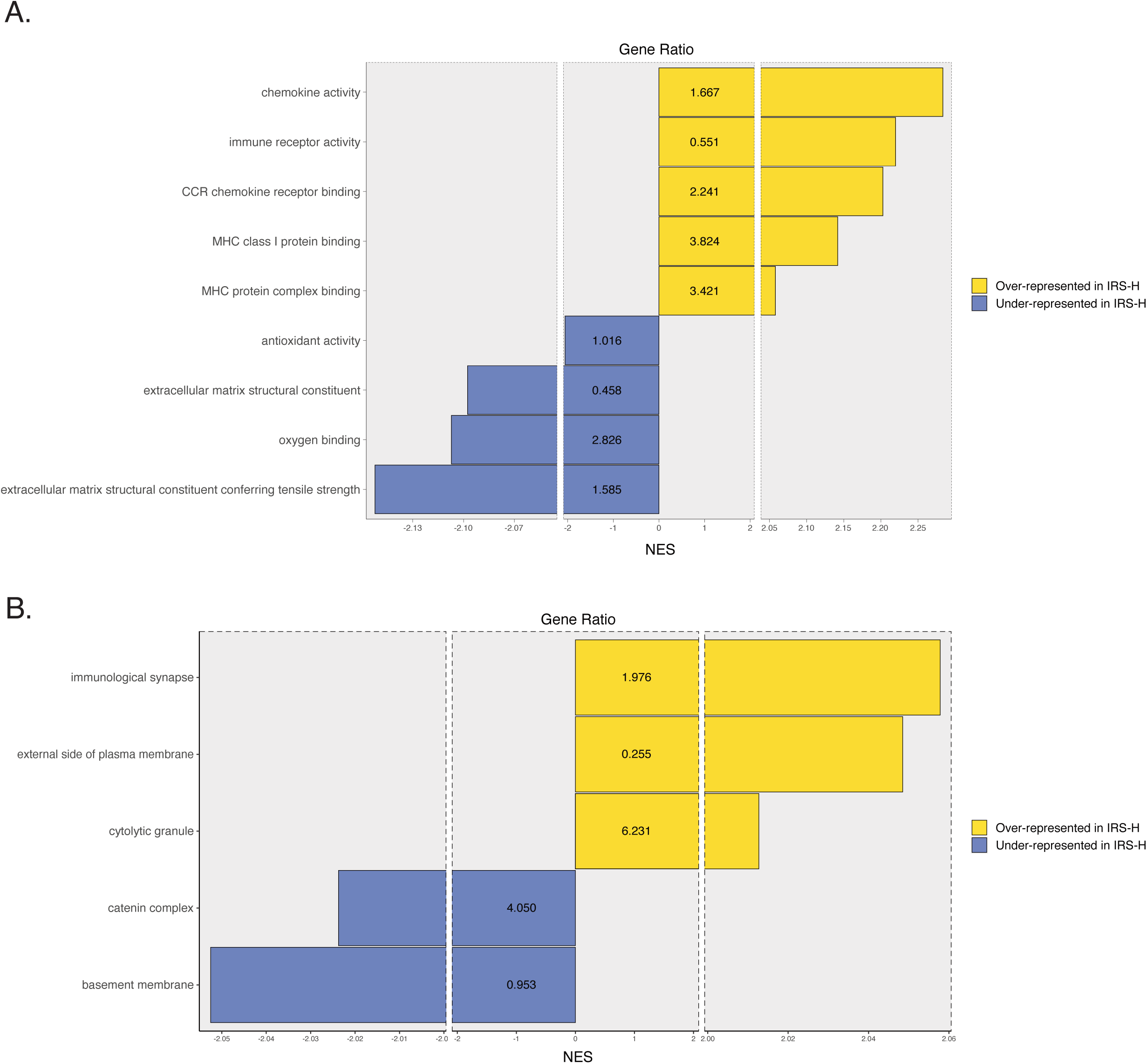
Gene set enrichment analysis showing statistically significantly over- or under-represented Gene Ontology **(A)** Molecular Functions and **(B)** Cellular Components in Immunotherapy Response Score (IRS) high (IRS-H) cases compared with IRS low (IRS-L) cases.

## Tables and titles

**Supplementary Table 1.** Efficacy endpoints.

| Endpoints | Results |
| --- | --- |
| **Response - no. (%)** |  |
| Complete response | 21 (8.05) |
| Partial response | 37 (14.18) |
| Stable disease | 43 (16.48) |
| Progressive disease | 126 (48.28) |
| Not evaluable | 34 (13.03) |
| **Overall response rate - % (95% CI)** | 22.22 (17.33 – 27.56) |
| **Disease control rate - % (95% CI)** | 38.70 (32.75 – 44.90) |
| **Median overall survival - months (95% CI)** | 8.90 (7.06 – 10.91) |

Abbreviations: CI, confidence interval.

**Supplementary Table 2.** Gene set enrichment analysis showing Gene Ontology data about **(A)** Biological Processes (BP), **(B)** Molecular Functions (MF), and **(C)** Cellular Components (CC) in Immunotherapy Response Score (IRS) high (IRS-H) cases compared with IRS low (IRS-L) cases.

